# Evaluation of the specificity and accuracy of SARS-CoV-2 rapid antigen self-tests compared to RT-PCR from 1015 asymptomatic volunteers

**DOI:** 10.1101/2022.02.11.22270873

**Authors:** Thomas Iftner, Angelika Iftner, Diana Pohle, Peter Martus

## Abstract

**Objective:** Evaluation of the specificity and accuracy of four CE-approved SARS-CoV-2 antigen rapid self-tests (AG-ST) Anbio, Clungene, Hotgene and Mexacare.

**Method:** 1015 asymptomatic volunteers were screened for SARS-CoV-2 by means of an oropharyngeal swab taken by qualified personnel and subsequent RT-PCR testing. Each participant additionally performed nasal self-swabs for two of the four rapid antigen tests at the same day according to the manufacturers’ instructions. Study participants transmitted a photo and own interpretation of their test results to the study center. The results of the two self-tests provided by the participants were correlated with the results of the SARS-CoV-2 RT-PCR and independently assessed and evaluated by the study center.

**Results:** None of the volunteers tested positive upon RT-PCR, whereas 13 AG-ST showed a false positive test result (0.7 %). The highest false positivity rate was found for the Clungene test (2.1 % compared to 0.2 % for the other tests), while the highest test failure rate (invalid) was found for the Mexacare test (3.7%). The Anbio and Hotgene tests produced the fewest false positive results when evaluated by the participants and also showed the best agreement among themselves.

**Conclusion:** SARS-CoV-2 Antigen rapid self-tests with higher false positive test rates, such as the Clungene test, or with high rates of invalid test results, such as the Mexacare test, are less suitable for screening purposes of asymptomatic study participants especially in low-prevalence settings. False positive or inadequate test results increase the burden on certified test laboratories due to verification PCR tests and cause a substantial economic loss due to unnecessary quarantine measurements and cause psychological stress in the affected study participants. In addition to earlier defined requirements for sensitivity for SARS-CoV-2 detection, a lower acceptance boundary for the false positivity rate of < 0.3% should be demanded.

## Introduction

Numerous rapid antigen tests for self-testing (AG-ST) to detect an infection with SARS-CoV-2 are available on the European market. Most of them are based on lateral flow immunochromatography and target the viral nucleoprotein (N) in respiratory samples, or, rarely, the spike protein (S). A number of different SARS-CoV-2 variants, including variants of concern (VOC) and variants of interest (VOI) have been transmitted worldwide. Since mutations primarily affect the S and ORF1a/b genes (1, 2), the vast majority of SARS-CoV-2 AG-ST using the N-antigen for detection are unaffected by these genetic changes. RT-PCR or transcription-mediated amplification (TMA) are the gold standard for SARS-CoV-2 detection. With a high level of sensitivity and specificity, they are able to detect SARS-CoV-2 at very early and very late stages of infection (3, 4). Nevertheless, as high viral loads are usually present during the early phase of viral infection in nasal and oropharyngeal samples, especially for the latest VOCs delta and omicron (5-7), certified AG-ST represent an important instrument in the control of the pandemic outbreak of SARS-CoV-2 and its variants.

The German state of Baden-Wuerttemberg (BW) has been distributing up to 3.5 million self-tests per week from various manufacturers since April 2021. For some of these self-tests, a conspicuously high number of false-positive results has sporadically been self-reported by the users, especially from primary and secondary schools. A self-test conducted under the guidance of or supervision by a trained third party will immediately result in the quarantine of the tested person and requires a confirmation via RT-PCR. In contrast, a positive result of a non-supervised self-test does not lead to quarantine, but establishes an obligation to retest by means of PCR diagnostics. Thus, false-positive results have far-reaching consequences with regard to the obligation to quarantine and lead to an additional burden on PCR capacities. However, data are scarce on the performance of these AG self-tests in real life scenarios.

In the European Union (EU), regulatory requirements for SARS-CoV-2 *in vitro* diagnostic (IVD) devices are defined in the IVD Directive 98/79/EC (IVD) (8) and such devices must have a certification (CE label) issued by the manufacturer before being allowed to be distributed at the EU common market. Exceptions are however possible because of the high dynamics of the pandemic situation as by comparison to the performance of a similar test for professional use such as the same antigen test for medical professionals. As AG-STs allow the rapid identification of acutely infected individuals by self-testing and thereby help to interrupt infection chains, they facilitate quick measures such as containment, isolation of patients in hospitals, and prevention of infection events via Health Care Workers (HCW). In addition, AG-STs can help to save limited reagents for RT-qPCR or TMA in diagnostic labs for symptomatic individuals (9, 10).

In a recent publication by Scheiblauer and colleagues (11), the sensitivities of different AG tests were determined for 122 CE-labeled antigen tests with a defined limit of a minimum sensitivity of 75 % in samples with Cycle threshold (Ct) values less than 25 cycles using a panel of diagnostic samples that had been collected by medical professionals and had been analyzed by SARS-CoV-2 RT-PCR before. This requirement was fullfilled by 96/122 tests, including tests from Teda (Anbio), Hotgen, and Clongene. Others, such as Mexacare’s test (sensitivity of 52.9 %) did not reach the sensitivity criterion. The data suggest a good suitability of a number of tests, but should be considered critically due to missing confidence intervals for the reported sensitivities. Data on the accuracy and especially on the specificity of self-taken nasal swab antigen tests under real-life conditions compared to a gold standard such as RT-qPCR are, however, scarce.

We therefore attempted to collect those data with 1015 asymptomatic volunteers that participated in a study, where an oropharyngeal swab was taken by qualified personnel and subsequently analyzed for the presence of SARS-CoV-2 by an RT-PCR test. Each participant additionally performed two consecutive nasal self-swabs for two rapid antigen tests at the same day (randomly chosen from the 4 BfArM listed AG-STs (12) published at PEI webpage (13) Anbio, Clungene, Hotgene and Mexacare) according to the manufacturer’ s instructions and transmitted a photo of the test results to the study center together with information on the self-judged test result. The results of the two self-tests provided by the participants were first correlated with the results of the SARS-CoV-2 RT-PCR and secondly independently assessed and evaluated by the study center.

## Materials and Methods

Four different SARS-CoV-2 rapid antigen self-tests (AG-ST; Table 1) were analyzed in an everyday use setting for their accuracy and compared to Altona RealStar RT-PCR as gold standard.

**Table 1.** Number of individual tests.

| Test name | Manufacturer, designation, article no. and BfArM no. | Issued | Missing* | For Evaluation |
| --- | --- | --- | --- | --- |
| **Anbio** | Teda Laukoetter Technologie GmbH<br>"COVID-19 Antigen Rapid Test (colloidal gold)<br>ANBIO Corona Antigen Nasal Swab"<br>Article no. C-10013b and C-10013c<br>BfArM no. 5640-S-079/21 | 505 | 37 | 468 |
| **Clungene** | Hangzhou Clongene Biotech Co., Ltd.<br>"COVID-19 Antigen Rapid Test" box of 5<br>BfArM no. 5640-S-168/21 | 512 | 33 | 479 |
| **Hotgen** | Beijing Hotgen Biotech Co., Ltd.<br>"Corona Lay Test, Home Self-Test. Coronavirus<br>(2019-nCoV) Antigen Test"<br>BfArM no. GZ 5640-S-057/21 | 502 | 38 | 464 |
| **Mexacare** | Mexacare GmbH<br>"MEXACARE Corona Home-Test "Antigen"<br>Rapid Test for self-testing"<br>Article no. 3211003<br>BfArM no. 5640-S-049/21 | 511 | 20 | 481 |
| **TOTAL** |  | **2030** | **128** | **1892** |

\* Tests without response, not performed and/or without indication of results by the test persons

For this, 1015 asymptomatic employees (unknown vaccination status) of the University Hospital Tübingen, Germany, who voluntarily agreed to participate in the study (ethical approval 391/2021BO2) were tested completely anonymous for SARS-CoV-2 by means of an oropharyngeal swab taken by qualified personnel and subsequent RT-PCR testing using the Altona RealStar kit (performed in a laboratory certified according to DIN EN ISO standard 15189). SARS-CoV-2 RT-PCR results were communicated to the study participant together with the respective barcode by encrypted e-mail. Each participant additionally performed nasal self-swabs without supervision for two of the four randomly selected rapid antigen tests on the day of oropharyngeal swab collection according to the manufacturers’ instructions (the order was determined by the participant) and sent a photograph of the test results by means of a bar-coded form without participant’s name via e-mail to an e-mail address specifically set up for the study.

138 test results could not be considered for further evaluation because 67 study participants did not provide a response (67 * 2 = 134 tests) (Table 1). In addition, one test could not be performed (lack of fluid), one subject did not send in a photograph of the test, and one subject did not self-report the test result on the form (2 tests).

The self-reported results were independently assessed and evaluated by Medical Virology staff in a Delphi procedure using the transmitted photographs of the test results to identify evaluation and/or application errors by the study participants. The anonymized results of the AG-ST of the volunteers together with the examiners’ results and the RT-PCR results were statistically analyzed by the Institute of Clinical Epidemiology and Applied Biometry (University Hospital Tuebingen).

Statistical analysis was performed by using SPSS (program release 26). Comparative analysis of the sensitivity of the AG-ST was not possible due to the very low prevalence of SARS-CoV-2 at the time of the study (May 12^th^ 2021 to July 20^th^ 2021) as all SARS-CoV-2 RT-PCR tests were negative. Comparative analysis of the false positivity rate, test accuracy and rate of invalid test results of the four AG-STs investigated was performed.

## Results

### Evaluation of the self-reported test results

In 97.4 % (1843/1892) of the test results, the study participants indicated a negative test result, in 0.7 % (13/1892), the test result was reported as “positive”, and in 1.9 % (36/1892), the result was reported as “unclear” (Table 2). An evaluation of the results revealed significant differences in the false positivity rates and the respective percentages of unclear test results between different AG-ST systems. With 2.1 %, the false positive rate was statistically significantly higher for the AG-ST Clungene compared to the three other AG-ST Anbio, Hotgen and Mexacare, each of which had a false positive rate of 0.2% (p = 0.007, Chi-square test). On the other hand, the lack of readability of the test result (unclear or invalid) was highest for the Mexacare test (3.7 % of cases). The differences of Mexacare versus Anbio (1.1 % unclear, p = 0.008) and Clungene (0.6 % unclear, p = 0.001) were statistically significant, the difference versus Hotgen was in the range of random variation (2.2 %, p = 0.152).

**Table 2.** Self-reported results of the different AG-ST rapid tests.

|  |  | Negative | Positive | Unclear | Total |
| --- | --- | --- | --- | --- | --- |
| **Anbio** | Number | 462 | 1 | 5 | 468 |
|  | % | 98.7 | 0.2 | 1.1 | 100.0 |
| **Clungene** | Number | 466 | 10 | 3 | 479 |
|  | % | 97.3 | 2.1 | 0.6 | 100.0 |
| **Hotgen** | Number | 453 | 1 | 10 | 464 |
|  | % | 97.6 | 0.2 | 2.2 | 100.0 |
| **Mexacare** | Number | 462 | 1 | 18 | 481 |
|  | % | 96.0 | 0.2 | 3.7 | 100.0 |

To better assess the suitability for the “real-life setting” for each AG-ST, the number of false positive results reported by the study participants was weighted twice, and the number of unclear cases was weighted once to obtain a combined comparison parameter (Table 3). The lower this score, the fewer false positives and unclear results have been produced by the respective self-test. By using this weighting score, the two tests Mexacare and Clungene performed worse than Anbio and Hotgen; Anbio performed statistically significantly better than Clungene (p = 0.036) and Mexacare (p = 0.026). The other differences were within the range of random variation.

**Table 3.** Number of false-positive and unclear AG-ST (determined by study participants) per test. Calculation of a performance score indicating the suitability in the “real life setting” (low value = high suitability).

|  | False-positive |  | Unclear |  | Score |
| --- | --- | --- | --- | --- | --- |
|  | Cases | % | Cases | % |  |
| **Anbio** | 1/468 | 0.2 | 5/468 | 1.1 | 7 |
| **Clungene** | 10/479 | 2.1 | 3/479 | 0.6 | 23 |
| **Hotgen** | 1/464 | 0.2 | 10/464 | 2.2 | 12 |
| **Mexacare** | 1/481 | 0.2 | 18/481 | 3.7 | 20 |

### Re-evaluation of the self-reported test results of the study participants by staff examiners

After re-evaluating of self-reported tests by staff examiners, the statistical analysis showed a concordance between the result of the study participants and the examiners of 99.8 % for a “negative test result”, 69.2 % for “positive”, and 86.1 % for “unclear” (Table 4). The random-corrected kappa measure for overall agreement, commonly used in medical statistics, was 86.8 %. From this analysis, it appears that only the Anbio and Hotgene test may be suitable for testing of asymptomatic study participants.

**Table 4.**
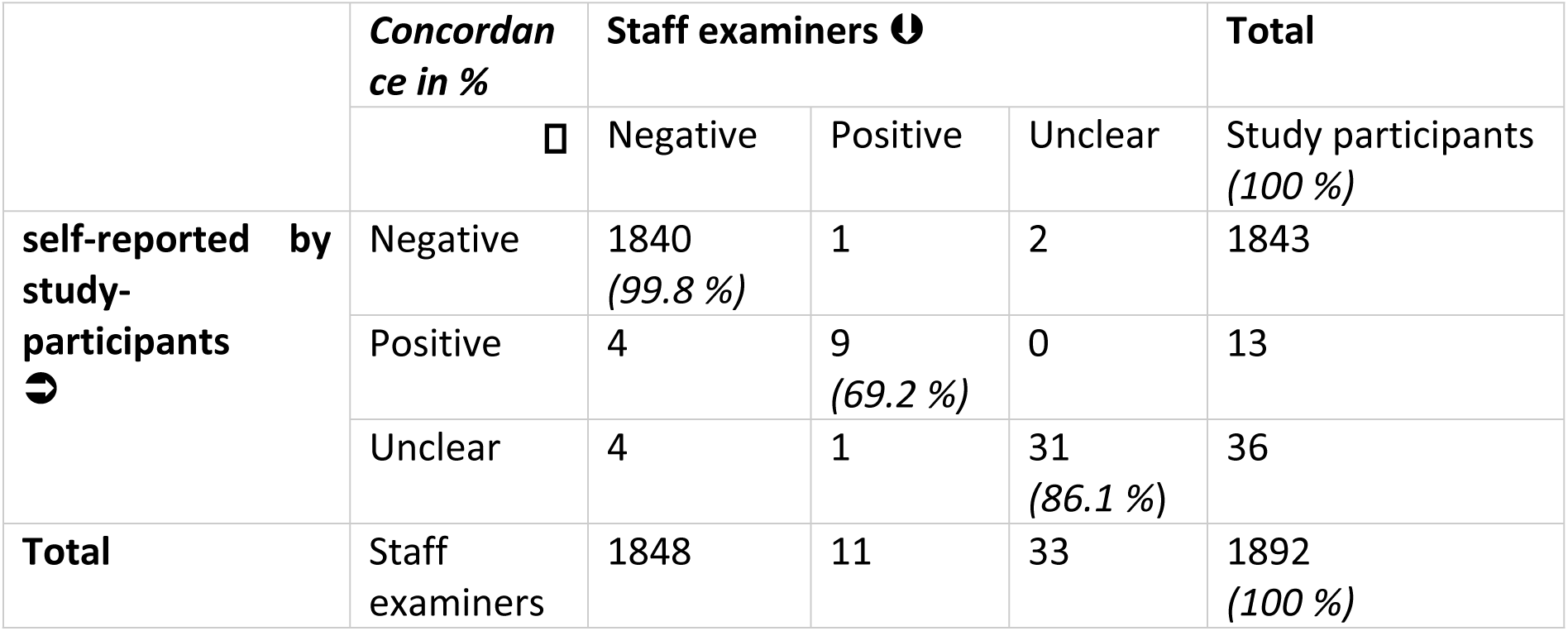
Results of the re-evaluation of self-reported AG-ST by staff examiners.

|  | **Concordance in %** | **Staff examiners** |  |  | **Total** |
| --- | --- | --- | --- | --- | --- |
|  |  | Negative | Positive | Unclear | Study participants (100 %) |
| **self-reported by study-participants** | Negative | 1840 (99.8 %) | 1 | 2 | 1843 |
|  | Positive | 4 | 9 (69.2 %) | 0 | 13 |
|  | Unclear | 4 | 1 | 31 (86.1 %) | 36 |
| **Total** | Staff examiners | 1848 | 11 | 33 | 1892 (100 %) |

Concordance was calculated for all test results together and again separately for all four AG-ST used (Table 5). For the four AG-ST rapid tests examined, an over 99.8 % concordance for a negative test result between self-reported results and results obtained by staff examiners was found.

**Table 5.** Results of the evaluation of the individual AG-ST by study participants and staff examiners.

|  | test result | study participant<br>A | examiner<br>B* | concordance |  |
| --- | --- | --- | --- | --- | --- |
|  |  |  |  | C | C:A |
|  |  |  |  | in cases | in % |
| **Anbio** | negative | 462 | 464 | 461 | 99.8 |
|  | positive | 1 | 1 | 0 | 0.0 |
|  | unclear | 5 | 3 | 3 | 60.0 |
| **Clungene** | negative | 466 | 468 | 465 | 99.8 |
|  | positive | 10 | 10 | 9 | 90.0 |
|  | unclear | 3 | 2 | 1 | 33.3 |
| **Hotgen** | negative | 453 | 454 | 452 | 99.8 |
|  | positive | 1 | 0 | 0 | 0.0 |
|  | unclear | 10 | 10 | 9 | 90.0 |
| **Mexacare** | negative | 462 | 464 | 462 | 100.0 |
|  | positive | 1 | 0 | 0 | 0.0 |
|  | unclear | 18 | 18 | 18 | 100.0 |

\*The numbers achieved by the examiners were higher than the number by self-reports due to missing information provided by some study participants.

The number of false positive rapid test results was highest for Clungene with 9 (concordance self-report and examiner = 90 %). The test Anbio, Hotgene and Mexacare each provided only one false positive result each, according to the self-testing person. However, this could not be confirmed during review by staff examiners. The highest number of unclear test results was observed with the AG-ST of Mexacare (18 cases) and Hotgene (10 cases by self-report and staff examiner). In contrast, the test result “unclear” was less frequent with the AG-ST Anbio and Clungene.

The test results “unclear” self-reported by the study participants were analyzed in more detail by the examiners (Table 6). While the study participants could only select between three choices to indicate the test result (“negative”, “positive” and “unclear”), the examiners distinguished between “negative”, “positive”, “unclear” and “invalid”. 4/36 tests self-reported as “unclear” were evaluated by the examiner as “negative” and 1/36 tests as “positive”. One test result was also classified as “unclear” by the examiners. The majority of the “unclear” subject test results were, however, rated as “invalid” by the examiners because the control band was not visible on the test strip. With 18 cases, this occurred most frequently in the AG-ST Mexacare.

**Table 6.** Re-evaluation of the self-reported test results by the examiners.

|  | Study participant | Staff examiner |  |  |  |
| --- | --- | --- | --- | --- | --- |
| | Unclear | Unclear | Negative | Positive | Invalid [% of $\Sigma$] |
| **Anbio** | 5 |  | 2 |  | 3 [3 %] |
| **Clungene** | 3 |  | 1 | 1 | 1 [1 %] |
| **Hotgen** | 10 | 1 | 1 |  | 8 [27 %] |
| **Mexacare** | 18 |  |  |  | 18 [60 %] |
| **Total $\Sigma$** | **36** | **1** | **4** | **1** | **30 [100 %]** |

Finally, study participants and staff examiners both listed problems or errors associated with the performance or evaluation of the tests (Table 7). While a missing control band was reported in 18 cases, lack of test buffer, shifted or illegible detection areas were also reported.

**Table 7.**
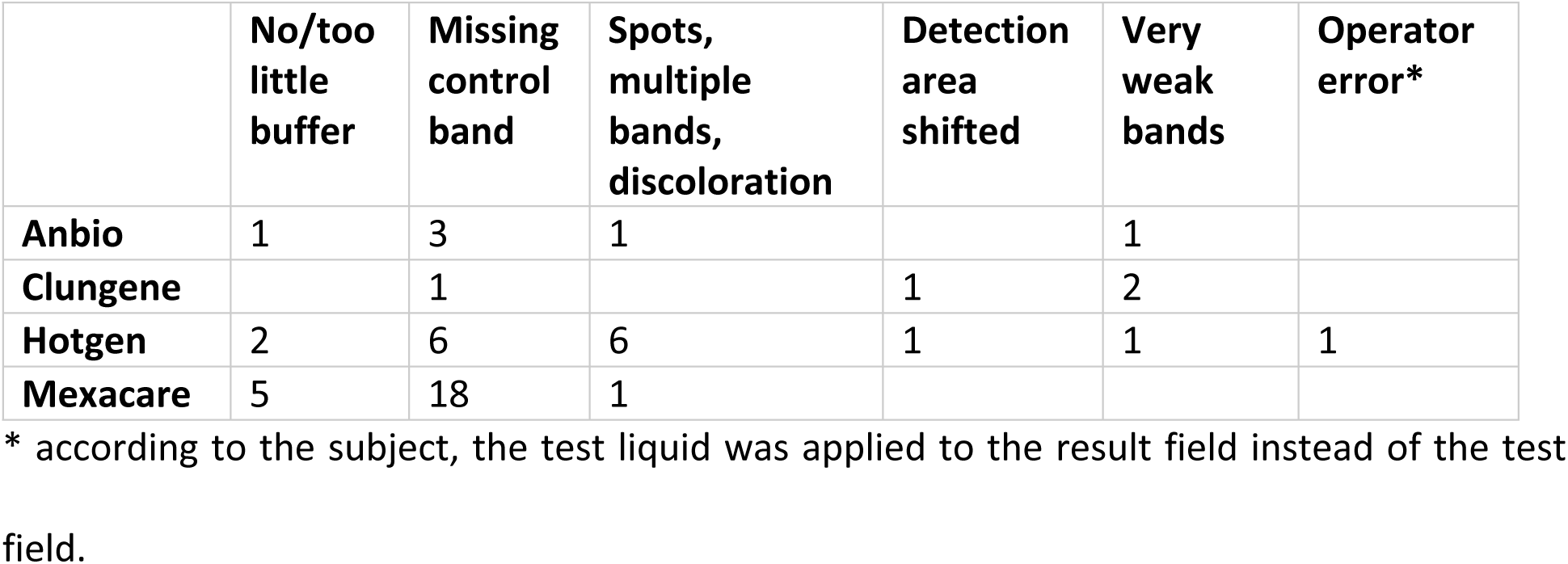
AG self-test problems or errors observed by study participants and staff.

## Discussion

There is a clear correlation between the infectivity of SARS-CoV-2 and high viral loads in aerosols produced by acutely infected persons, independent of the presence or absence of symptoms (14-16). Lateral flow immunoassays, commonly designated as rapid antigen tests (AG), are known to have a limited sensitivity, but are able to detect high viral loads in respiratory specimens. Thus, they are also commonly used for screening of asymptomatic individuals to detect infections to limit the spread of infection. For example, SARS-CoV-2 AG rapid self-tests (AG-ST) are used as part of a self-testing strategy in order to avoid that health-care workers infected with SARS-CoV2 infect vulnerable groups at work. A large number of different AG-ST detecting the nuclear antigen of all known variants of SARS-CoV-2 are commercially available. Almost all of them state sensitivity values of > 92% and specificities of 99.1% for PCR-confirmed specimens in their self-declared performance data, with which they would meet the quality requirements: sensitivity > 80 % for PCR-pos. and specificity 97 % in a study population ≥ 100 persons (13). However, these self-declared performance data are often incorrect as recently shown in a study for test sensitivity of 122 CE-marked AG-ST (11). In the study presented here, we wanted to investigate now the specificity of such tests and evaluate the frequency of handling and interpretation errors due to usage by non-professionals.

By screening 1015 asymptomatic volunteers for SARS-CoV-2 RNA and comparing this to AG-ST results, we could not confirm the self-reported specificity for one of four AG-ST tests, while specificity was acceptable for the other tree tests. In addition, some AG-ST may have an unacceptable high level of test result drop outs, e.g. due to illegible detection areas or missing control bands. In our present study, the highest false positivity rate was 2.1 % for the Clungene test, and the highest test failure rate (invalid) was 3.7 % for the Mexacare test. The Anbio and Hotgene tests produced the fewest false positives when used by study participants and also showed the best agreement among themselves. As none of the participants had a confirmed SARS-CoV-2 infection (i.e. RT-PCR was negative in all cases), it was not possible to investigate the sensitivity of the AG-ST. This was likely due to the very low SARS-CoV-2 prevalence in summer 2021. A potential limitation of the study might be a relatively high number of medical staff, e.g. participants likely were better trained to use AG-ST as the rest of the general population. Therefore, handling or interpretation errors could be underestimated in this cohort. As additional criteria might affect the performance of AG-ST it would have been useful to allow the user to describe in a free text field of the form sheet used to transfer the test results to the study center the possible source of interpretation errors, e.g. by lack of readability of the test, missing components in the kits etc.

In summary, our data show that the Clungene and Mexacare tests are probably less suitable for usage as antigen self-tests due to their high rate of false positive results and high number of unclear/invalid results, respectively. Tests with a high false-positivity rate should especially be avoided to be used in a population with a low prevalence of SARS-CoV-2.

## Data Availability

The data underlying the results presented in the study are available from (Prof. Peter Martus)

## Acknowledgements

We thank all participants enrolled in the study. We acknowledge A. Nothacker for organizing and conducting the clinical part and A. L. Bissinger for assistance with study protocol and ethics approval and Tina Ganzenmueller for critical reading of the manuscript. This work was supported by “Ministerium für Soziales, Gesundheit und Integration Baden-Württemberg”. We acknowledge support by Open Access Publishing Fund of University of Tübingen.

## Notes

### Competing Interest Statement

The authors have declared no competing interest.

### Funding Statement

The funders had no role in study design, data collection and analysis, decision to publish, or preparation of the manuscript.

### Author Declarations

University hospital Tübingen, Medical Faculty, Ethics Commitee. ethical approval nr. 391/2021BO2 declared that no written consent necessary.

